# ChatGPT and the Clinical Informatics Board Examination: The End of Knowledge-based Medical Board Maintenance?

**DOI:** 10.1101/2023.04.25.23289105

**Authors:** Yaa Kumah-Crystal, Scott Mankowitz, Peter Embi, Christoph U. Lehmann

## Abstract

**Objectives:** Assess ChatGPT’s performance on the Clinical Informatics Board Examination (CIBE) and discuss the implications of large language models (LLMs) for board certification and maintenance.

**Materials and Methods:** We tested ChatGPT using 260 multiple-choice questions from Mankowitz’s Clinical Informatics Board Review book, omitting six image-dependent questions.

**Results:** ChatGPT answered 190 (74%) of 254 eligible questions correctly. While performance varied across the Clinical Informatics Core Content Areas, differences were not statistically significant.

**Discussion:** ChatGPT’s performance raises concerns about the potential misuse in medical certification and the future validity of knowledge assessment exams. While ChatGPT is able to answer multiple-choice questions accurately, relying on AI systems for exams will compromise the credibility and validity of at-home assessments and undermine public trust.

**Conclusion:** The advent of AI and LLMs threatens to upend existing processes to board certification and maintenance and necessitates new approaches to the evaluation of proficiency in medical education.

## Background and Significance

The use of large language models (LLMs) such as OpenAI’s ChatGPT^[1]^ has shown promise answering knowledge questions and passing exams as demonstrated by its recent success passing the United States Medical Licensing Examination (USMLE) exams with a 60% grade^[2]^. The Clinical Informatics Board Exam’s (CIBE) focus and content differ from the USMLE^[3]^. Unlike the USMLE, which tests knowledge and its application in basic sciences and clinical medicine, the CIBE assesses knowledge and its application in health information technology^[4]^. As clinical informatics focuses on improving patient care and outcomes, which require an application of concepts and principles, the CIBE may present unique challenges for LLMs. Furthermore, as board certification exams move towards self-paced, self-administered assessments, the availability of tools such as ChatGPT raises concerns about the consequences of exam-takers using such resources to pass maintenance exams and its effect on credentialing organizations to accurately evaluate an individual’s level of expertise and proficiency^[5]^.

## Objectives

Our study assessed ChatGPT’s ability to pass practice exams for the CIBE and its performance in the core competencies of the CIBE. We discuss the potential implications of using LLMs for board certification and maintenance.

## Materials and Methods

We used a corpus of 260 multiple-choice questions from Mankowitz’s Clinical Informatics Board Review book published in 2018^[6]^. The questions represent the knowledge areas tested on the examination administered by the American Board of Preventive Medicine. Questions were categorized according to the Core Content for the Subspecialty of Clinical Informatics^[7 8]^. Questions depending on the use of images were omitted. Each question was entered into ChatGPT 3.5 with a brief preamble requesting justification why the answer suggested by ChatGPT was correct. The question was considered answered correctly, if ChatGPT could identify the answer that correlated to the book’s answer key.

## Results

Of 260 questions, six (3%) were excluded because they relied on visual stimuli to deliver the context, leaving 254 (97%) questions available for analysis. Of the remaining 254 questions, ChatGPT answered 190 (74%) correctly. Categorized based on the Clinical Informatics Core Content Areas schema, ChatGPT performed from best to worst in 1. Fundamental Knowledge and Skills (85%), 2. Leadership and Professionalism (76%), 3. Data Governance and Data Analytics (74%), 4. Enterprise Information Systems (72%), and 5. Improving Care Delivery and Outcomes (71%). However, chi-square analysis did not reveal any statistically significant differences in ChatGPT’s performance across these categories (X^2^(4) = 0.59, p = 0.96). (Table 1)

**Table 1:** ChatGPT Performance on Categories of CIBE Questions

| Clinical Informatics Category | Correct/Total |
| --- | --- |
| Fundamental Knowledge and Skills | 28/33 (85%) |
| Leadership and Professionalism | 52/68 (76%) |
| Data Governance and Data Analytics | 17/23 (74%) |
| Enterprise Information Systems | 28/39 (72%) |
| Improving Care Delivery and Outcomes | 65/91 (71%) |
| **Total** | **190/254 (75%)** |

## Discussion

Our study demonstrated that ChatGPT has the ability to answer multiple-choice questions with a high degree of accuracy, achieving a passing score expected of fellowship-trained informaticians on the CIBE. However, the prevalence of at-home board certification and maintenance of certification (MOC) examinations raises concerns about the potential (mis)use of freely available tools like ChatGPT in professional medical certification, calling into question the validity of these knowledge assessment exams and, by extension, risk undermining the public’s trust in board certification in general.

While ChatGPT could potentially aid an individual in passing the MOC examination, it should not be relied upon by life-long learners. The fundamental purpose of MOC is to reinforce important concepts and principles and allow takers an understanding of their skills and knowledge^[9]^. While the use of ChatGPT during an MOC exam could encourage the individual to think more critically about these concepts as they answer questions, this approach also carries a risk of reducing critical reflection. With open-book MOC exams that currently allow access to primary texts and search engines like Google, individuals are still required to assimilate information to determine the correct answer^[10]^. However, using LLMs to obtain answers to multiple-choice questions requires no familiarity with the theory and its applications as the answers are provided directly.

## Limitations

Our study was limited by the fact that the sample of questions used in the study was derived from a single source, Mankowitz’s Clinical Informatics Board Review book, which may not represent the full range of question types and content encountered on the actual CIBE. Additionally, questions that contained images could not be evaluated. Incorporating more pictographs into exams could prevent questions from being answered by today’s LLMs. However, this is unlikely to be a permanent solution as newer GPT models are more proficient at interpreting images. In the future, organizations that certify may prohibit the use of LLMs and will introduce novel kinds of questions designed to identify individuals who use LLMs in MOC exams.

## Conclusion

As the availability and usage of LLMs continue to expand and as ChatGPT-3.5 is already being replaced by GPT-4, contemplating the significance of LLMs for medical education and for the validity of board certification exams is imperative. While our study demonstrated that ChatGPT has the capacity to answer multiple-choice questions with a high degree of accuracy, it is not advisable to rely solely on AI systems for exam-taking purposes due to the potential risk of compromising the credibility and validity of at-home assessments. Although it is possible for physicians to uphold ethical standards and engage with LLMs to promote personal growth and skill development, our findings cast doubt that an answer easily obtained through an LLM can truly serve as an accurate assessment of an individual’s proficiency in the subject matter^[11 12]^. LLMs will force the development of new approaches to the evaluation and measurement of mastery. Alternatively, they may force certifying organizations to revert to proctored, in-person exams^[10]^. Ultimately, the advent of AI necessitates a corresponding evolution in education to adapt to this new environment.

## Data Availability

All data produced in the present study are available upon reasonable request to the authors

## Acknowledgments

We express our gratitude to Sydney Roth for her valuable data analysis and editing assistance, which greatly enhanced the paper’s quality and clarity.

## References

1. OpenAI. ChatGPT [Internet]. https://openai.com/blog/chatgpt/ (accessed Feb 1, 2023).

2. Kung TH, Cheatham M, Medenilla A, et al. Performance of ChatGPT on USMLE: Potential for AI-assisted medical education using large language models. PLOS Digit Health 2023;2(2):e0000198.

3. United States Medical Licensing Examination [Internet]. https://www.usmle.org/ (accessed Feb 1, 2023).

4. Chan KS, Zary N. Applications and Challenges of Implementing Artificial Intelligence in Medical Education: Integrative Review. JMIR Med Educ 2019;5(1):e13930.

5. Clinical Informatics [Internet]. https://www.theabpm.org/become-certified/subspecialties/clinical-informatics/ (accessed Feb 1, 2023).

6. Mankowitz S. Clinical Informatics Board Review and Self Assessment. Springer International Publishing; 2018 Feb 8

7. Gardner RM, Overhage JM, Steen EB, et al. Core content for the subspecialty of clinical informatics. J Am Med Inform Assoc 2009;16(2):153–7.

8. Silverman HD, Steen EB, Carpenito JN, et al. Domains, tasks, and knowledge for clinical informatics subspecialty practice: results of a practice analysis. J Am Med Inform Assoc 2019;26(7):586–93.

9. Hawkins RE, Lipner RS, Ham HP, et al. American Board of Medical Specialties Maintenance of Certification: theory and evidence regarding the current framework. J Contin Educ Health Prof 2013;33 Suppl 1:S7–19.

10. Zagury-Orly I, Durning SJ. Assessing open-book examination in medical education: The time is now. Med Teach 2021;43(8):972–73.

11. Topol EJ. High-performance medicine: the convergence of human and artificial intelligence. Nat Med 2019;25(1):44–56.

12. Liévin V, Hother CE, Winther O. Can large language models reason about medical questions?. arXiv preprint arXiv:2207.08143. 2022 Jul 17.

